# *GIPC1* intermediate-length repeat expansion in amyotrophic lateral sclerosis

**DOI:** 10.1101/2025.05.22.25328088

**Authors:** Kohei Muto, Konoka Tachibana, Ryosuke Miyamoto, Yuki Kuwano, Naoki Kihara, Hiroki Yamazaki, Yusuke Osaki, Masaki Kamada, Seishu Banzai, Hiroki Ueno, Tatsuya Fukumoto, Nazere Keyoumu, Naoko Matsui, Koji Fujita, Masahiro Nakamori, Yu Yamazaki, Hirofumi Maruyama, Yuishin Izumi, Hiroyuki Morino

## Abstract

Repeat expansion diseases, particularly those involving GC-rich motifs, have been increasingly recognized as contributors to neurological and neuromuscular disorders. Amyotrophic lateral sclerosis (ALS) has been linked to several such expansions, including intermediate-length repeats in genes implicated in oculopharyngodistal myopathy (OPDM). To investigate the possible involvement of CGG repeat expansions in ALS, 424 ALS patients and 312 controls of Japanese descent were screened for expansions in five genes associated with repeat expansion disorders, namely *GIPC1*, *RILPL1*, *FMR1*, *AFF2*, and *NUTM2BAS1*. Repeat-primed PCR and fragment analysis revealed that four ALS patients exhibited abnormal CGG expansions in *GIPC1* (33–55 repeats), whereas two control individuals harbored expanded alleles (67 and 83 repeats). No expansions in the other genes were detected. Long-read sequencing confirmed repeat sizes and showed sequence instability. Histopathological analysis of ALS patients with *GIPC1* expansion demonstrated classical ALS pathology, including phosphorylated TDP-43-positive inclusions. RNA fluorescence in situ hybridization revealed nuclear foci containing *GIPC1* repeat RNA exclusively in ALS patients with *GIPC1* expansions, suggesting RNA-mediated toxicity. These findings indicate that a subset of ALS patients present with intermediate CGG expansions in *GIPC1*, which may represent a novel pathogenic mechanism analogous to other noncoding repeat disorders. Given that *GIPC1* full expansions are associated with OPDM, these results support the hypothesis of a pathological continuum between neurodegeneration and myopathy driven by repeat length and sequence context. Nonetheless, further investigations into the potential of *GIPC1* CGG expansions as genetic risk factors or modifiers in ALS are warranted.

## Introduction

Advances in long-read sequencing technologies have markedly accelerated the discovery of repeat expansion diseases. To date, approximately 60 repeat expansion diseases, predominantly neurological and neuromuscular, have been identified.^1^ However, data from the 100,000 Genomes Project suggest that the actual detectable prevalence of repeat expansion diseases may be two- to three-fold higher than currently recognized,^2,3^ implying the existence of numerous undetected pathogenic repeat expansions underlying neurological disorders.

Amyotrophic lateral sclerosis (ALS) is a progressive neurodegenerative disorder characterized by the selective degeneration of upper and lower motor neurons, leading to muscle atrophy, paralysis, and ultimately respiratory failure.^4^ Aside from the well-established hexanucleotide repeat expansion in the *C9orf72* gene (MIM: 614260), repeat expansion mechanisms in several other genes have been implicated in ALS.^5,6^ Notably, studies have shown that intermediate-length expansions in *ATXN2* (MIM: 601517), *LRP12* (MIM: 618299), and *NOTCH2NLC* (MIM: 618025) were associated with increased ALS risk.^7–9^ In contrast, full-length expansions in *LRP12* and *NOTCH2NLC* have been associated with oculopharyngodistal myopathy (OPDM), a disorder that presents with ptosis, dysphagia, and distal limb muscle weakness.^10^ These observations suggest that repeat length may modulate the clinical phenotype, with shorter expansions promoting neurodegeneration and longer expansions causing myopathy.

To date, five genes have been associated with OPDM, namely *LRP12*, *GIPC1* (MIM: 605072), *NOTCH2NLC*, *RILPL1* (MIM: 614092), and more recently *ABCD3* (MIM: 170995), each of which is characterized by either CGG or CCG repeat expansion in the 5′ untranslated region (5′ UTR).^10–13^ This remarkable convergence on GC-rich repeat motifs, regardless of gene function, highlights the importance of the repeat sequence in driving disease development.^10,14^ The fact that *LRP12* and *NOTCH2NLC* are implicated in both ALS and OPDM indicates that these two conditions may represent different clinical manifestations along a shared pathological spectrum. This spectrum is likely defined by the presence and instability of GC-rich repeat expansions, with repeat length and sequence context determining whether the disease presents with neurodegenerative or myopathic features.

Therefore, the current study aimed to investigate intermediate CGG repeat expansions in several genes previously implicated in neurological and neuromuscular disorders, including OPDM and fragile X syndrome (FXS; MIM: 300624), within a cohort of ALS patients.

## Materials and methods

### Subjects

This study enrolled a total of 424 patients clinically diagnosed with ALS, including 7 individuals with a positive family history and 1 patient with spinocerebellar degeneration (SCD) without known causative repeat expansions as a disease control. Moreover, 312 individuals without neurologically disorders were included as controls. All participants were of Japanese descent. ALS was diagnosed by board-certified neurologists based on the revised El Escorial criteria for possible, probable, or definite ALS.^15^ Patients with pathogenic variants in genes known to be associated with ALS, such as *C9orf72*, *SOD1* (MIM: 147450), *FUS* (MIM: 137070), *TARDBP* (MIM: 605078), and *TBK1* (MIM: 604834), were excluded from this study. This study was approved by the Ethics Committee of Tokushima University (3261-11) and was conducted in accordance with the tenets of the Declaration of Helsinki. Written informed consent was obtained from all participants.

### DNA extraction

Genomic DNA was extracted from the peripheral blood of the participants using QuickGene-610L (Wako, Osaka, Japan). DNA concentration and purity were assessed using NanoDrop One (Thermo Fisher Scientific, Waltham, MA).

### Repeat-primed PCR (RP-PCR) and fragment analysis

We selected five genes in which aberrant CGG repeat expansions in the 5′ UTR have been implicated in neuromuscular diseases other than ALS. The associated phenotypes were as follows: *GIPC1* with OPDM2 (MIM: 618940)^11^; *RILPL1* with OPDM4 (MIM: 619790)^12^; *FMR1* (MIM: 309550) with FXS, fragile X-associated tremor/ataxia syndrome (FXTAS; MIM: 300623), and premature ovarian failure type 1 (POF1; MIM: 311360)^16^; *AFF2* (MIM: 300806) with X-linked intellectual developmental disorder-109 (XLID109; MIM: 309548)^17^; and *NUTM2BAS1* (MIM: 618639) with oculopharyngeal myopathy with leukoencephalopathy-1 (OPML1; MIM: 618637).^10^ Repeat expansions in these five genes were initially screened in 424 ALS patients using RP-PCR. Subsequently, fragment analysis was performed to determine the CGG repeat size of *GIPC1* in 424 ALS patients, 1 patient with SCD, and 312 healthy controls. A histogram was constructed based on the repeat length of the longer alleles in each individual determined through fragment analysis. The threshold for normal repeat size was set at 32 repeats or fewer based on both the distribution observed in our control group and prior studies suggesting pathogenicity above this threshold.^11^ Primers and PCR conditions for RP-PCR and fragment analysis are detailed in Tables S1 and S2. Electrophoresis was performed using SeqStudio Flex (Thermo Fisher Scientific) according to the manufacturer’s instructions. GeneScan (Thermo Fisher Scientific) was used to analyze fragment size data.

### Long-read sequencing with Cas9-mediated enrichment

Long-read sequencing was performed on four ALS samples with a CGG repeat expansion (ALS^exp+^-1 to ALS^exp+^-4), two ALS samples without an expansion (ALS^exp−^-1 and ALS^exp−^-2), two control samples with an expansion (NC^exp+^-1 and NC^exp+^-2), and one control sample without an expansion (NC^exp−^-1). The CGG repeat regions of *GIPC1* (GenBank: NM_005716.4) located in the 5′ UTR were selectively enriched using a CRISPR-Cas9-based target enrichment system. Guide RNA sequences for enrichment are listed in Table S3. Library preparation was performed using the SQK-LSK109 kit (Oxford Nanopore Technologies, Oxford, UK). Sequencing was conducted on the MinION platform (Oxford Nanopore Technologies) using FLO-MIN109 flow cells, following to the manufacturer’s protocol. Basecalling was performed using Dorado (version 0.7, https://github.com/nanoporetech/dorado). Sequencing reads were aligned to the human genome using LAST (version 1584, https://gitlab.com/mcfrith/last) and minimap2 (version 2.28, https://github.com/lh3/minimap2). Repeat length was estimated using NanoRepeat (version 1.8.1, https://github.com/WGLab/NanoRepeat), whereas consensus sequences of the repeat regions were generated using tandem-genotypes (version 1.9.2, https://github.com/mcfrith/tandem-genotypes). DNA 5-methylcytosine (5mC) methylation status was assessed using DeepMod2 (version 0.3.2, https://github.com/WGLab/DeepMod2). Haplotype phasing of reads was performed using WhatsHap (version 2.3, https://github.com/whatshap/whatshap). Methylation profiles across the repeat regions were visualized using the Integrative Genomics Viewer (version 2.19.3, https://igv.org/). Moreover, a waterfall plot of repeat sequences was generated using RepeatAnalysisTools (https://github.com/PacificBiosciences/apps-scripts.git).

### Statistical analysis

Differences in repeat size between the ALS and healthy control groups were evaluated using the Mann–Whitney U-test. Statical analysis were performed using R software (version 4.1.3, https://www.r-project.org/).

### Histology

Formalin-fixed, paraffin-embedded brain and spinal cord sections were obtained from autopsied patients (ALS^exp+^-2 and ALS^exp−^-3). Staining of 6-µm-thick sections was then performed using hematoxylin–eosin and Klüver–Barrera methods. Immunoreaction product deposits on immunohistochemically stained sections were visualized using a Histofine Simple Stain MAX-PO MULTI (Nichirei Biosciences, Tokyo, Japan), 3,3′-diaminobenzidine (Nichirei Biosciences), and primary antibodies against phosphorylated TDP-43 (phospho Ser409/410) (clone 11-9, mouse monoclonal, dilution 1:5000, Cosmo Bio Co Ltd, Tokyo, Japan), p62 (clone 3/P62 LCK LIGAND, mouse monoclonal, dilution 1:300, BD Biosciences, San Jose, CA), GIPC1 (rabbit polyclonal, dilution 1:200, Proteintech Group, Chicago, IL), and GIPC1 (clone BG-12, mouse monoclonal, dilution 1:200, Santa Cruz Biotechnologies, Santa Cruz, CA). Antigen retrieval was performed using heat within citrate buffer (pH 6.0). All slides were counterstained with hematoxylin to visualize neurons and other cells.

### RNA fluorescence in situ hybridization (FISH)

To investigate the potential association between RNA-mediated toxicity and CGG repeat expansions in *GIPC1*, RNA FISH was performed on spinal cord sections from the following three individuals: an ALS patient with *GIPC1* repeat expansion (ALS^exp+^-2); an ALS patient without an expansion (ALS^exp−^-3); and a SCD patient without an expansion (SCD^exp−^-1) as disease control. Two Cy5-labeled oligonucleotide probes were designed to detect *GIPC1* mRNA: one to the CGG repeat sequence [*GIPC1*-(CGG)_8_] and the other complementary to the downstream sequence (*GIPC1*-5′UTR) (Figure 1A). Formalin-fixed spinal cord sections from the patients were prehybridized in a solution containing 50% formamide, 10% dextran sulfate, 0.1 mg/ml tRNA, and 2× saline sodium citrate (SSC) for 1 h at 37 °C. Thereafter, sections were hybridized in a solution containing 50% formamide, 10% dextran sulfate, 0.1 mg/ml tRNA, 400 ng of Cy5-conjugated probes, and 2× SSC at 37 °C overnight. After hybridization, samples were washed twice with 50% formamide in 2× SSC at 37 °C for 20 min each, followed by two additional 20-min washes each with 2× SSC and 0.2× SSC at room temperature. Nuclei were then stained with Fixed Cell ReadyProbes reagent (DAPI) (Thermo Fisher Scientific), after which fluorescent images were acquired using a laser scanning confocal microscope (Nikon A1R, Nikon, Tokyo, Japan).

**Figure 1.**
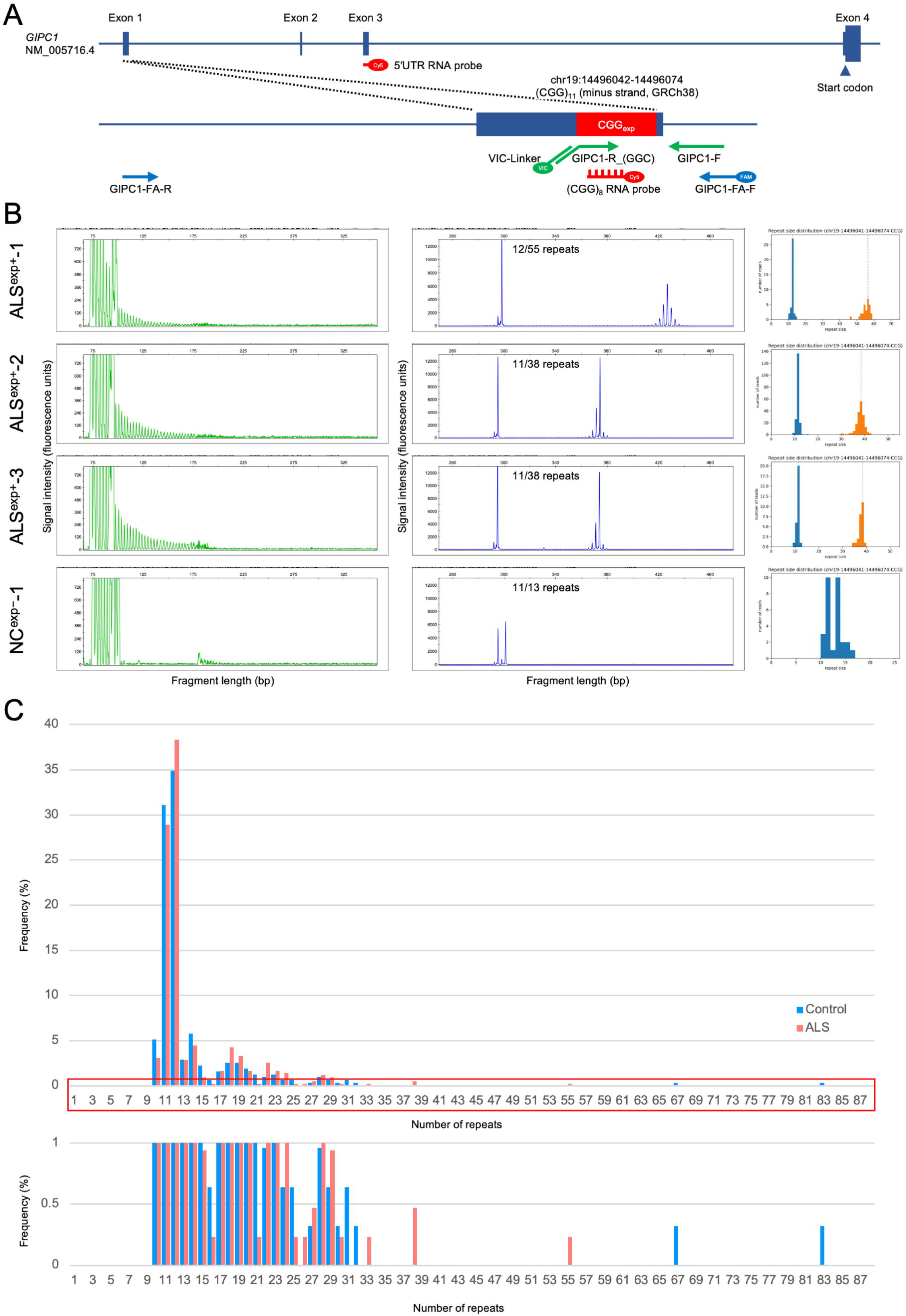
Analysis of CGG repeat expansions in the *GIPC1* gene. (A) The CGG repeat is located within exon 1 of *GIPC1*. Primers and probes for repeat-primed PCR (RP-PCR, green), fragment analysis (blue), and RNA fluorescence in situ hybridization (FISH, red) were designed within or adjacent to the repeat region. (B) The left panel shows the results for RP-PCR, in which abnormal CGG repeat expansions were detected in the upper three cases. The middle panel displays the results of fragment analysis. The right panel presents histograms of repeat sizes estimated from long-read sequencing data using NanoRepeat; orange bars indicate expanded alleles. (C) Histogram of CGG repeat sizes determined through fragment analysis. The lower graph shows an enlarged view of the low-frequency range. In the control group, most alleles contained 32 or fewer repeats. Two control samples exhibited abnormally expanded alleles, whereas four ALS samples harbored alleles with 33 or more repeats.

## Results

### Identification of abnormal CGG repeat expansions in GIPC1 among ALS patients

RP-PCR screening for *GIPC1* repeat expansions revealed a characteristic sawtooth tail pattern in eight ALS samples, indicating abnormal expansions (Figure 1B). In contrast, no abnormal repeat expansion patterns were observed in the other screened genes (*RILPL1*, *FMR1*, *AFF2*, and *NUTM2BAS1*). Subsequent fragment analysis confirmed abnormal CGG expansions exceeding the predefined threshold of 32 repeats in four ALS samples: ALS^exp+^-1 (55/12 repeats), ALS^exp+^-2 (38/11), ALS^exp+^-3 (38/11), ALS^exp+^-4 (33/12). Notably, two control samples, NC^exp+^-1 (83/12) and NC^exp+^-2 (67/10), also exhibited expanded alleles. Among the ALS samples with sawtooth RP-PCR patterns, four harbored repeat lengths below the cutoff (27–30 repeats). The CGG repeat size ranged from 10 to 55 in the ALS group and from 10 to 83 in controls (Figure 1C). Statistical analysis revealed no significant difference in repeat length distribution between the ALS and control groups (p = 0.7546).

Long-read sequencing analysis of the expanded alleles demonstrated that all four ALS samples with more than 32 repeats harbored uninterrupted, pure CGG tracts (Figure 2A). However, the two control individuals with expanded repeats exhibited complex interrupted sequences. In particular, NC^exp+^-1 carried a 5′ GGG substitution replacing the initial CAG, along with multiple AGG interruptions within the repeat region, whereas NC^exp+^-2 harbored an internal CAA interruption and a short deletion in the 5′ flanking region (Figure 2B). Such mutations in the 5′ flanking region were not detected in any of the ALS patients with repeat expansions. In all alleles with long CGG repeats, the first CAG motif at the 5′ end of the repeat region was uniformly replaced by CGG regardless of whether the samples were from ALS patients or controls. Analysis of DNA methylation status (5mC) around the *GIPC1* repeat region revealed no appreciable differences between ALS and control samples, regardless of expansion status (Figure S1).

**Figure 2.**
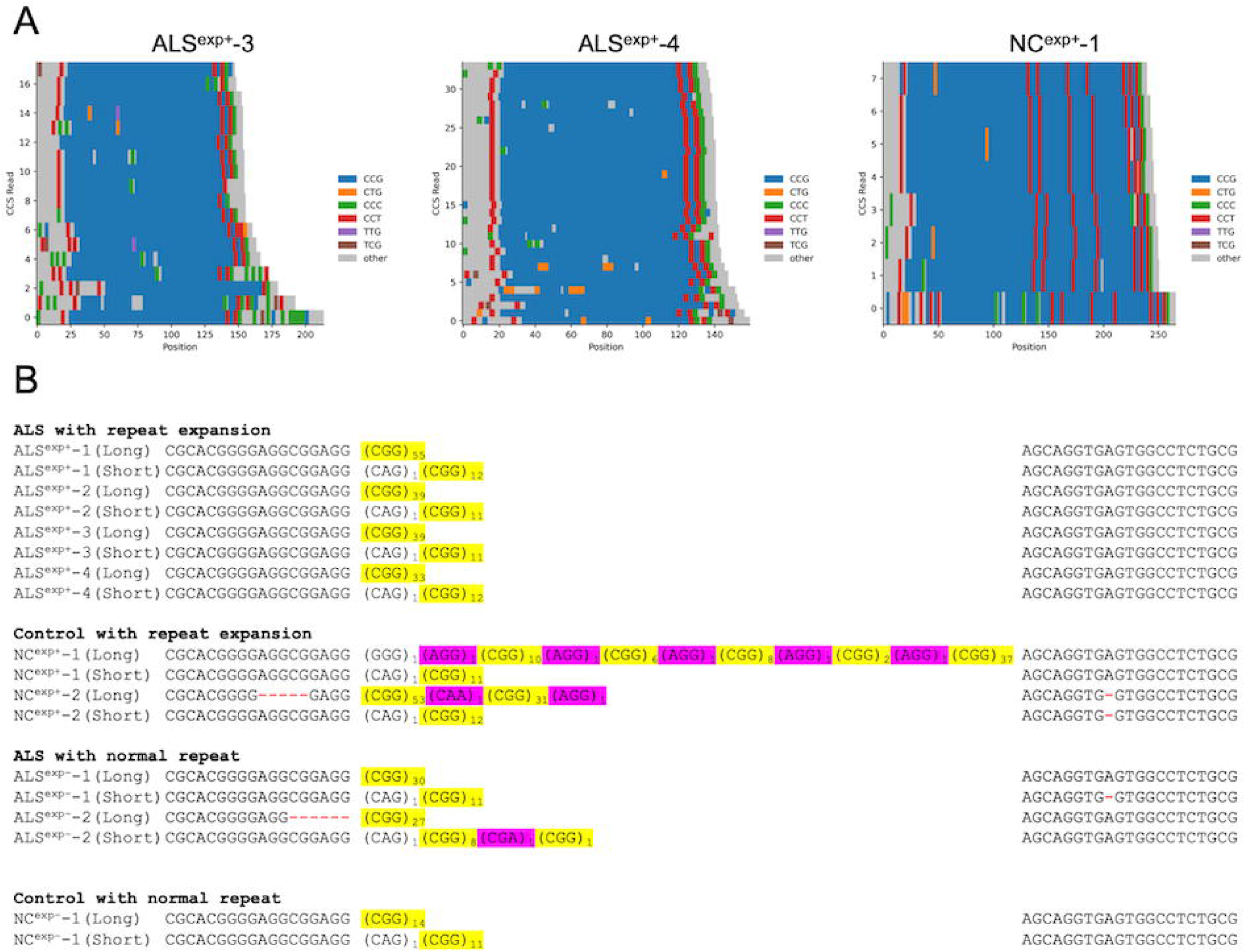
Structural features of CGG repeat expansions in *GIPC1* identified through long-read sequencing. (A) Waterfall plots of repeat sequences obtained by long-read sequencing demonstrated a uniformly expanded CGG repeat region in ALS samples. In contrast, control samples exhibited multiple interrupted sequences within the repeat region. (B) Consensus sequences generated from long-read data highlight repeat composition: yellow indicates CGG repeat tracts, whereas magenta denotes interrupted sequences. Deletions adjacent to the repeat region were also observed.

### Clinical characteristics

The mean age of ALS onset among the four patients harboring CGG expansions in *GIPC1* was 67 years (range, 50–75 years). The site of onset was the limb in three patients and both the limb and bulbar regions in one patient (Table 1). All patients exhibited both upper and lower motor neuron signs at the first clinical assessment. Disease duration from onset to death was 1, 6, and 7 years in the three patients, with one patient remaining alive 5 years after symptom onset. Three patients presented with classical ALS phenotype, whereas one exhibited a progressive bulbar phenotype. The patient with the longest disease duration of 7 years had the longest CGG repeat size (55/12). Although all deceased patients received noninvasive ventilation, none underwent tracheostomy with invasive mechanical throughout their disease course. Needle electromyography revealed no myogenic changes indicating OPDM in all four cases. None of the patients had a family history of ALS or neuromuscular disorders; however, the parent of ALS^exp+^-3 had been diagnosed with Parkinson’s disease. Three siblings of ALS^exp+^-3 were available for clinical and genetic evaluation, all of whom had no neurological abnormalities and carried an intermediate repeat expansion similar to the proband.

**Table 1.**
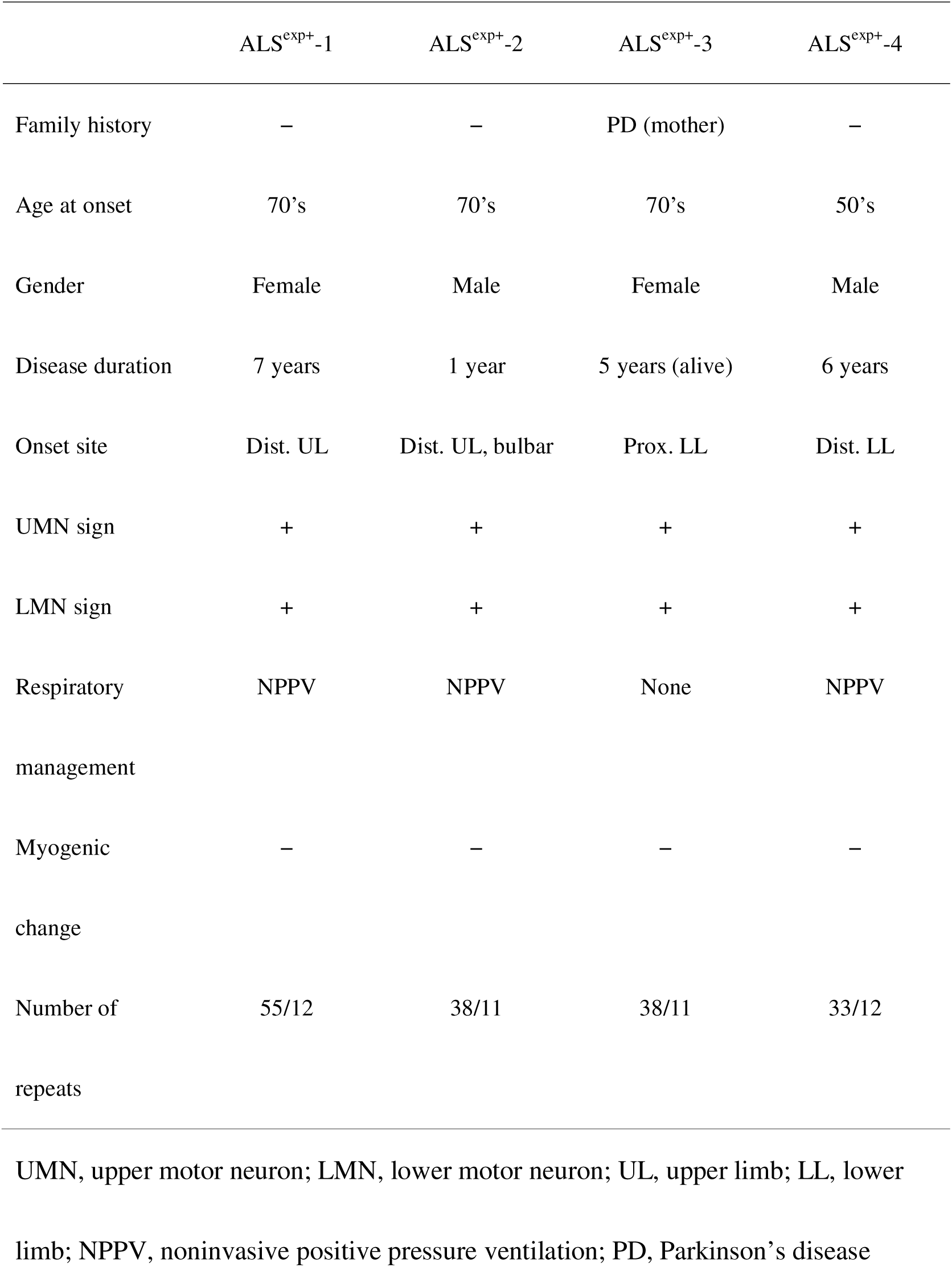
Clinical features of ALS patients with CGG repeat expansions in *GIPC1*.

|  | ALS<sup>exp+</sup>-1 | ALS<sup>exp+</sup>-2 | ALS<sup>exp+</sup>-3 | ALS<sup>exp+</sup>-4 |
| --- | --- | --- | --- | --- |
| Family history | – | – | PD (mother) | – |
| Age at onset | 70's | 70's | 70's | 50's |
| Gender | Female | Male | Female | Male |
| Disease duration | 7 years | 1 year | 5 years (alive) | 6 years |
| Onset site | Dist. UL | Dist. UL, bulbar | Prox. LL | Dist. LL |
| UMN sign | + | + | + | + |
| LMN sign | + | + | + | + |
| Respiratory management | NPPV | NPPV | None | NPPV |
| Myogenic change | – | – | – | – |
| Number of repeats | 55/12 | 38/11 | 38/11 | 33/12 |

UMN, upper motor neuron; LMN, lower motor neuron; UL, upper limb; LL, lower limb; NPPV, noninvasive positive pressure ventilation; PD, Parkinson's disease

### Postmortem neuropathological findings

Our findings showed neuronal loss in the anterior horn of the lumbar spinal cord and phosphorylated TDP-43-positive and p62-positive neuronal cytoplasmic inclusions. Similarly, neuronophagia and phosphorylated TDP-43-positive neuronal cytoplasmic inclusions were observed in the primary motor cortex, which were consistent with the neuropathological hallmarks of ALS (Figure 3). Furthermore, immunohistochemical staining of the spinal anterior horn using two different antibodies against GIPC1 revealed no abnormal structures with specific immunoreactivity (Figure S2).

**Figure 3.**
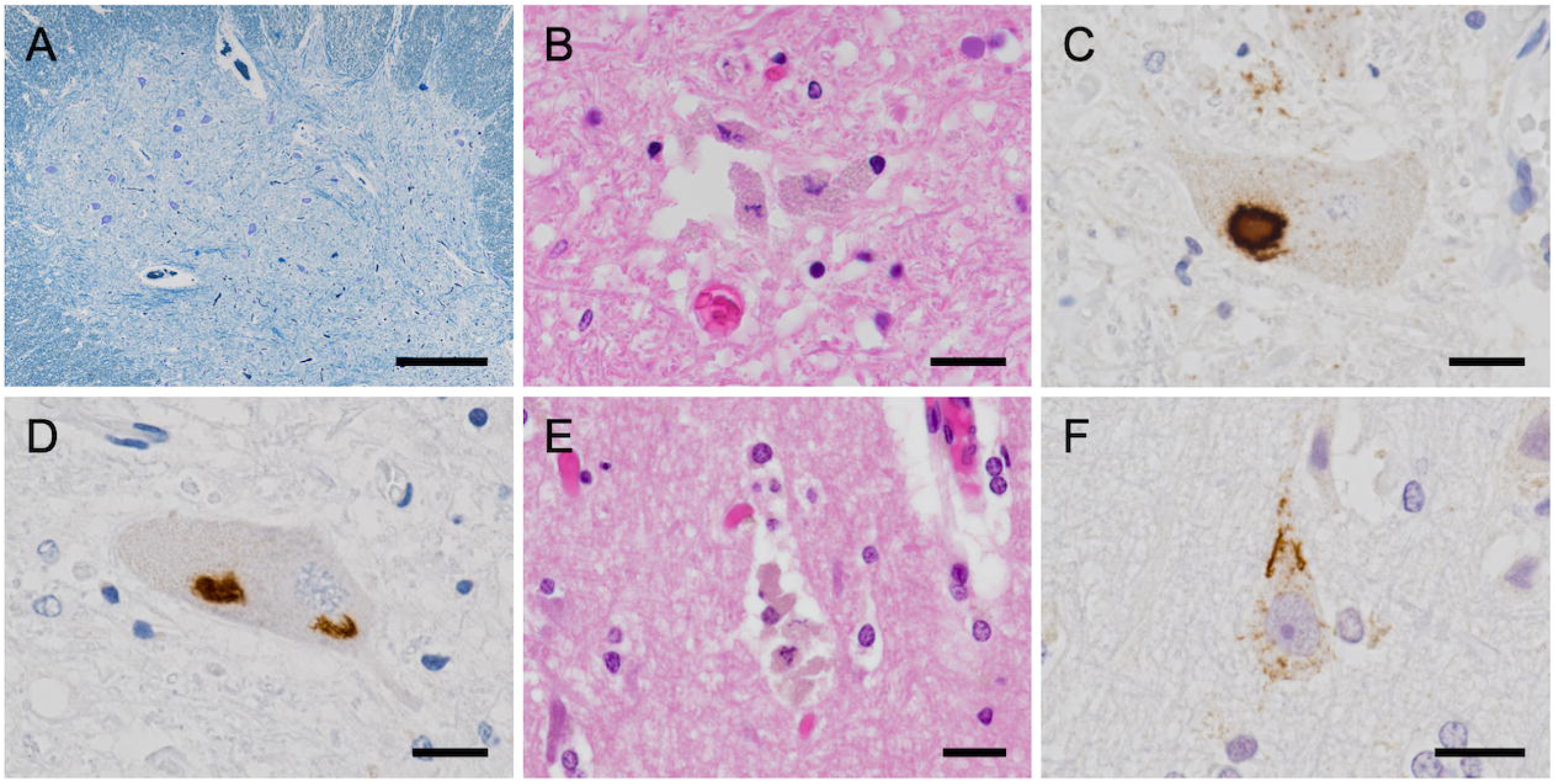
Neuropathological features in the ALS patient (ALS^exp+^-2) with *GIPC1* CGG repeat expansion. (A) Neuronal loss observed in the lumbar anterior horn. (B) Neuronophagia observed in the lumbar anterior horn. (C) Phosphorylated TDP-43-immunoreactive neuronal cytoplasmic inclusions observed in the lumbar anterior horn. (D) p62-immunoreactive neuronal cytoplasmic inclusions observed in the lumbar anterior horn. (E) Neuronophagia observed in the primary motor area. (F) Phosphorylated TDP-43-immunoreactive neuronal cytoplasmic inclusion observed in the primary motor area. Scale bars: (A) 500 μm; (B–F) 20 μm.

### RNA foci formation by repeat-expanded GIPC1 mRNA in ALS motor neurons

Intranuclear RNA foci formation of *GIPC1* mRNA containing CGG repeat expansions were observed in spinal cord neurons of the ALS patient with an expansion (ALS^exp+^-2) but not in either of the two control cases: the ALS patient without an expansion (ALS^exp−^-3) and the disease control with SCD (SCD^exp−^-1) (Figure 4). Foci were detected using probes targeting the CGG repeat and the 5′ untranslated region of *GIPC1* mRNA, indicating that these structures indicate the presence of repeat-containing *GIPC1* transcripts rather than nonspecific CGG-rich RNA. These findings suggest that nuclear accumulation of *GIPC1* mRNA with CGG repeat expansions may be implicated in the pathological process through which ALS developed in this particular case.

**Figure 4.**
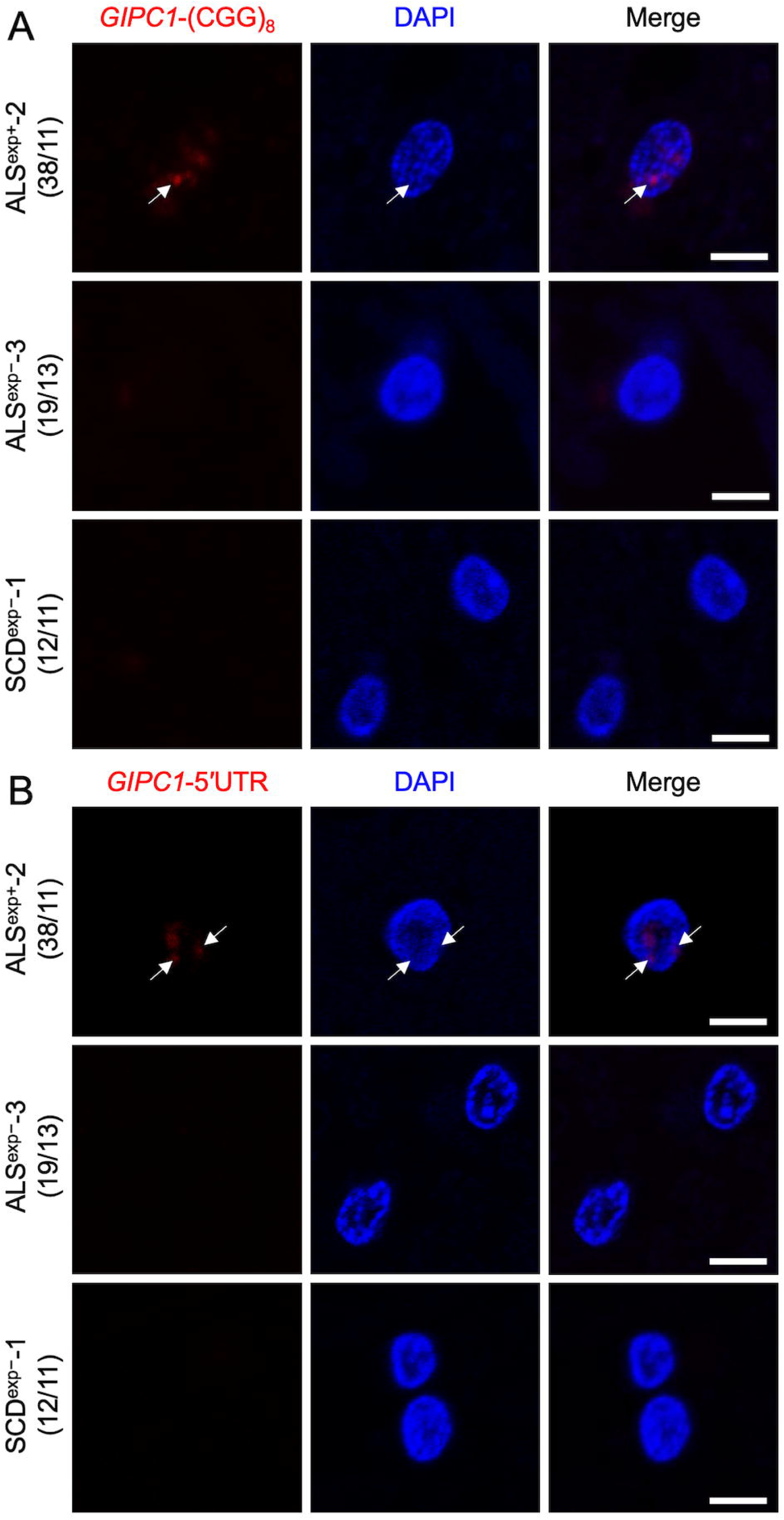
Detection of intranuclear RNA foci in the ALS patient with *GIPC1* CGG repeat expansion via RNA fluorescence in situ hybridization (FISH) (A) Results of RNA FISH using a (CGG)_8_ probe. Intranuclear RNA foci were observed exclusively in the ALS patient harboring a CGG repeat expansion in *GIPC1*. (B) RNA FISH results using another probe targeting the 5′ UTR of *GIPC1*. Similar intranuclear RNA foci were detected with this *GIPC1*-specific probe, confirming the presence of expanded repeat-containing RNA. Numbers in parentheses indicate the number of CGG repeats in *GIPC1*. Scale bars: 5 μm.

## Discussion

The present study identified a novel CGG repeat expansion in 5′ UTR of *GIPC1*, including TDP-43- and p62-positive cytoplasmic inclusions, among patients clinically and pathologically diagnosed with ALS. Although CGG repeat expansions in *GIPC1* have been associated with OPDM (*GIPC1*-OPDM),^18^ our findings suggest that intermediate-length expansions may instead induce a motor neuron phenotype, as observed in *LRP12* and *NOTCH2NLC*.^7,8,19^ Notably, RNA FISH using both (CGG)_8_ and *GIPC1*-specific probes demonstrated the formation of intranuclear RNA foci exclusively in the ALS patient harboring the repeat expansion, implicating the toxic RNA species in disease pathogenesis. The expanded alleles consisted of uninterrupted CGG repeats, with no difference in DNA methylation status at the repeat locus having been observed between the ALS and control samples. Collectively, these findings support the existence of an ALS phenotype associated with *GIPC1* repeat expansion (*GIPC1*-ALS).

In Western populations, GGGGCC repeat abnormalities in *C9orf72* have been the most prevalent genetic etiology for familial ALS,^20^ accounting for over 30% of such cases.^21–23^ In contrast, East Asian populations, including Japan, have a substantially lower frequency of *C9orf72*-ALS, accounting for only 2.3% of ALS cases.^21^ This low prevalence partially explains the higher proportion of genetically unidentified ALS cases in East Asia.^22^ The present study found that 0.94% of ALS cases, all of whom had no family history, presented with abnormal CGG repeat expansions in the *GIPC1* gene, indicating its contribution to sporadic ALS. This frequency is comparable to that of *FUS*-ALS,^22^ the second most common genetically defined ALS subtype in East Asia. Notably, full-length CGG repeat expansions in *LRP12*,^10^ *GIPC1*,^18^ *NOTCH2NLC*,^24^ and *RIPL1*^12^ have been established as causes of OPDM and together account for approximately 75% of genetically confirmed OPDM cases among Asians.^12^ These OPDM-related mutations, except for *ABCD3*,^13^ have not been observed in Western populations, suggesting a distinct ethnic distribution in the prevalence of CGG repeat expansion disorders. Given that intermediate-length expansions can predispose individuals to neurodegeneration,^2,25,26^ the higher frequency of such alleles among Asian populations may explain both the elevated incidence of OPDM and the potential contribution of OPDM-related gens to sporadic ALS among Asians.

RNA FISH performed in the current study detected RNA foci in spinal motor neurons of ALS patients with a CGG repeat expansion in *GIPC1*. GC-rich repeat motifs, such as CGG and GGGGCC, increase the susceptibility to the formation of stable secondary structures, including hairpins and G-quadruplexes,^27–30^ which can aggregate into insoluble RNA foci.^31,32^ Studies suggest that RNA foci acquire toxicity by sequestering RNA-binding proteins and disrupt essential cellular factors,^33,34^ contributing to ALS pathogenesis.^35^ Similar RNA foci have been consistently identified in *C9orf72*-ALS/FTD and FXTAS, with their number and distribution correlating with disease severity.^36–38^ Experimental studies have shown that expanded repeat RNA alone is sufficient to induce neurodegeneration.^39–41^ In our study, TDP-43 pathology was evident in autopsy samples that showed no accumulation of *GIPC1*-derived polypeptides, suggesting that RNA-mediated gain-of-function toxicity was the primary pathogenic mechanism.

Repeat expansion disorders often show phenotypic variability depending on the repeat length. For example, studies have shown that CGG expansions in *FMR1* cause FXS at full expansion and FXTAS at intermediate lengths.^16,42^ Identical repeat motifs in different genes can also produce similar phenotypes, as observed in familial adult myoclonus epilepsy (MIM: 601068), which has been associated with TTTCA repeat insertions in several genes.^14,43^ These findings suggest that the repeat motif itself, rather than the function of the host gene, may be the primary determinant of disease phenotype. In support of this concept, our findings suggest that CGG or CCG repeat motifs themselves, rather than the specific host gene, may play an important role in ALS pathogenesis.

Abnormal CGG repeat expansions in *GIPC1* were also observed in two individuals from the control group. In both cases, the repeats contained sequence interruptions and were accompanied by small indels in the 5′ flanking region, which may have mitigated the pathogenic potential of the expansions. Interruption within repeat tracts have been known to reduce RNA structural stability and attenuate toxicity and exert protective effects against several repeat expansion disorders, including spinocerebellar ataxias type 1 (SCA1)^44,45^ and type 10 (SCA10),^46^ myotonic dystrophy,^47–49^ Huntington’s desease,^50^ and FXTAS.^51,52^ Moreover, the 5′ flanking region plays a critical role in transcriptional regulation.^53–55^ The indels observed in this region may have decreased the expression of repeat-containing transcripts, thereby further reducing pathogenicity.

All ALS cases with *GIPC1* CGG repeat expansions were sporadic. However, one patient (ALS^exp+^-3) had asymptomatic siblings carrying the same expansions, suggesting low penetrance. Co-segregation could not be demonstrated in any family, implying that additional genetic, epigenetic, or environmental factors may have influenced disease manifestation. Further studies involving larger familial cohorts and multicenter collaborations are needed to clarify the pathogenic role and inheritance patterns of *GIPC1*-ALS.

In summary, our findings indicate that CGG repeat expansions, including those in *GIPC1*, may contribute to the pathogenesis of ALS through RNA-mediated toxicity as evidenced by the presence of RNA foci in affected motor neurons. The detection of such foci underscores the pathogenic potential of repeat RNA independent of host gene function. Moreover, the identification of repeat interruptions and flanking region indels in unaffected individuals suggests that repeat configuration modulates penetrance and toxicity. Although all identified cases were sporadic with no demonstrable familial co-segregation, our results suggest the presence of a previously unrecognized ALS subgroup defined by repeat structure. Further investigations using diverse patient cohorts and experimental models will be essential to clarify disease mechanisms and facilitate repeat-based molecular classification and personalized therapeutic strategies in ALS.

## Supporting information

Graphical Abstract

## Data Availability

The datasets generated during this study are available within the paper and in the supplemental information.

## Acknowledgments

We would like to acknowledge the affected patients and their families for their participation in this study. We thank Ms. Yoshimi Mihara for her excellent technical assistance. We also thank Dr. Tomoyasu Matsubara for his expert advice on pathological studies. This work was supported by MHLW Research on rare and intractable diseases Program Grant Number JPMH23FC1008. This work was also supported by the Serika Fund.

## Author contributions

Conceptualization, H. Morino, R.M., and K.M.; genetic analysis, K.M., K.T., R.M., H. Morino, N. Keyoumu, and M.K.; statistical analysis, K.T., R.M., and H. Morino; biochemical analysis, Y.K.; histopathological analysis, S.B.; clinical and diagnostic evaluation, K.M., R.M., N. Kihara, H.Y., Y.O., T.F., N.M., K.F., Y.I., H.U., M.N., Y.Y., and H. Maruyama; writing original draft, K.M., R.M., and H. Morino; review and editing, all authors.

## Web resources

The URLs for data presented herein are as follows:

Online Mendelian Inheritance in Man (OMIM), http://www.omim.org/ UCSC Genome Browser, http://genome.ucsc.edu

---

**Figure S1. Methylation status around the *GIPC1* CGG repeat region in ALS and control samples** Methylation profiles surrounding the CGG repeat region in the *GIPC1* gene, visualized using Integrative Genomics Viewer. Methylation data were obtained through Nanopore sequencing. The analyzed samples include four ALS patients with a CGG repeat expansion, one ALS patient without an expansion, one healthy control with an expansion, and one healthy control without an expansion. Red indicates regions of high methylation, whereas blue represents regions of low methylation.

**Figure S2. GIPC1 immunostaining of spinal cord sections from patients with ALS**

(A) Anterior horn of the lumbar spinal cord from an ALS patient with a CGG repeat expansion stained with anti-GIPC1 antibody (Proteintech).

(B) Anterior horn of the lumbar spinal cord from an ALS patient without a CGG repeat expansion stained with anti-GIPC1 antibody (Proteintech).

(C) Anterior horn of the lumbar spinal cord from an ALS patient with a CGG repeat expansion stained with anti-GIPC1 antibody (Santa Cruz Biotechnology).

(D) Anterior horn of the lumbar spinal cord from an ALS patient without a CGG repeat expansion stained with anti-GIPC1 antibody (Santa Cruz Biotechnology).

No significant GIPC1 aggregates or differences in staining patterns were observed between groups. Scale bar: 50 µm (A–D).

