## Supplementary figures and images for "*GIPC1* intermediate-length repeat expansion in amyotrophic lateral sclerosis"

### Graphical Abstract

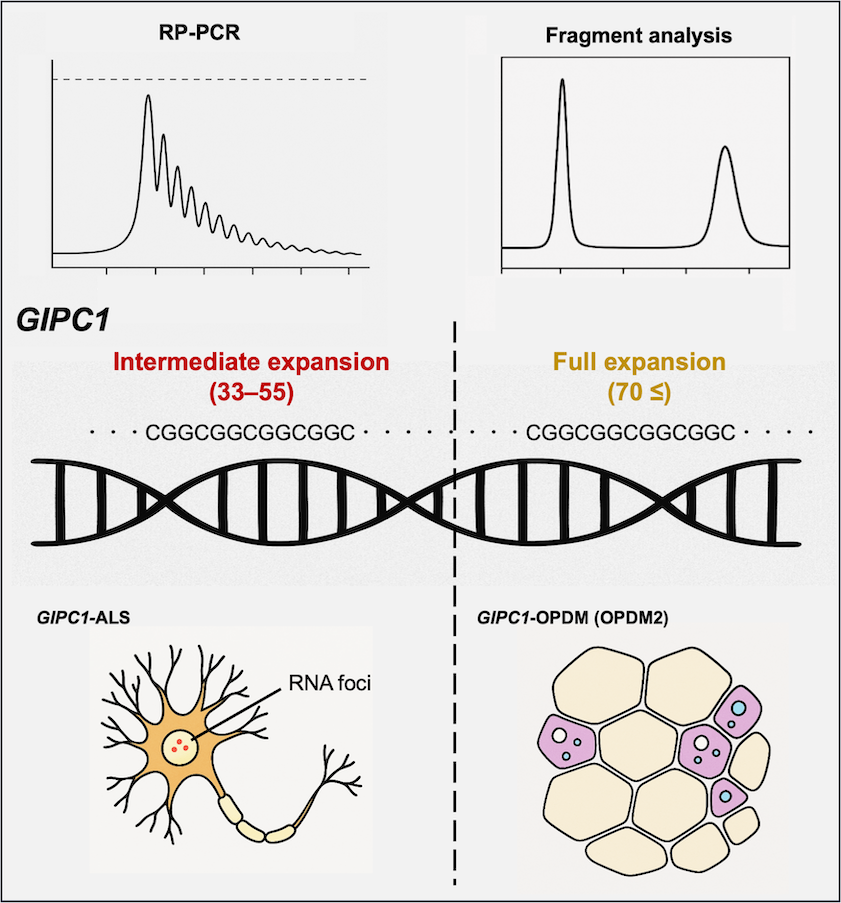

### Supplemental Figure 1

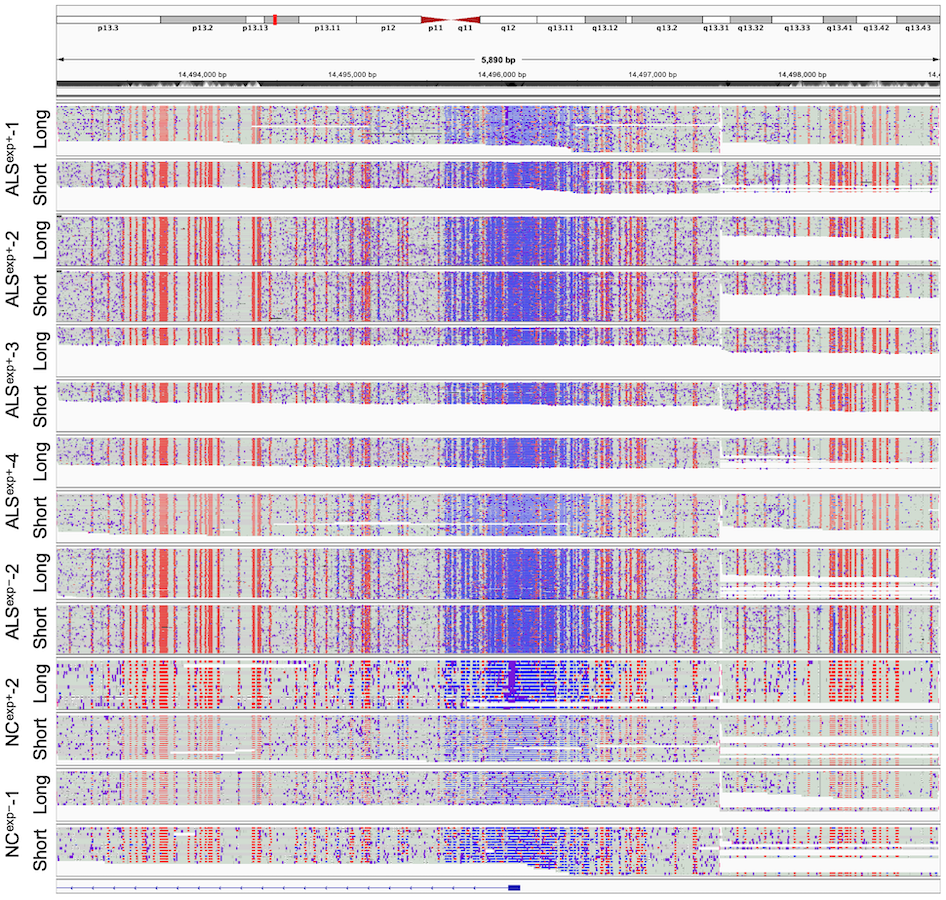

### Supplemental Figure 2

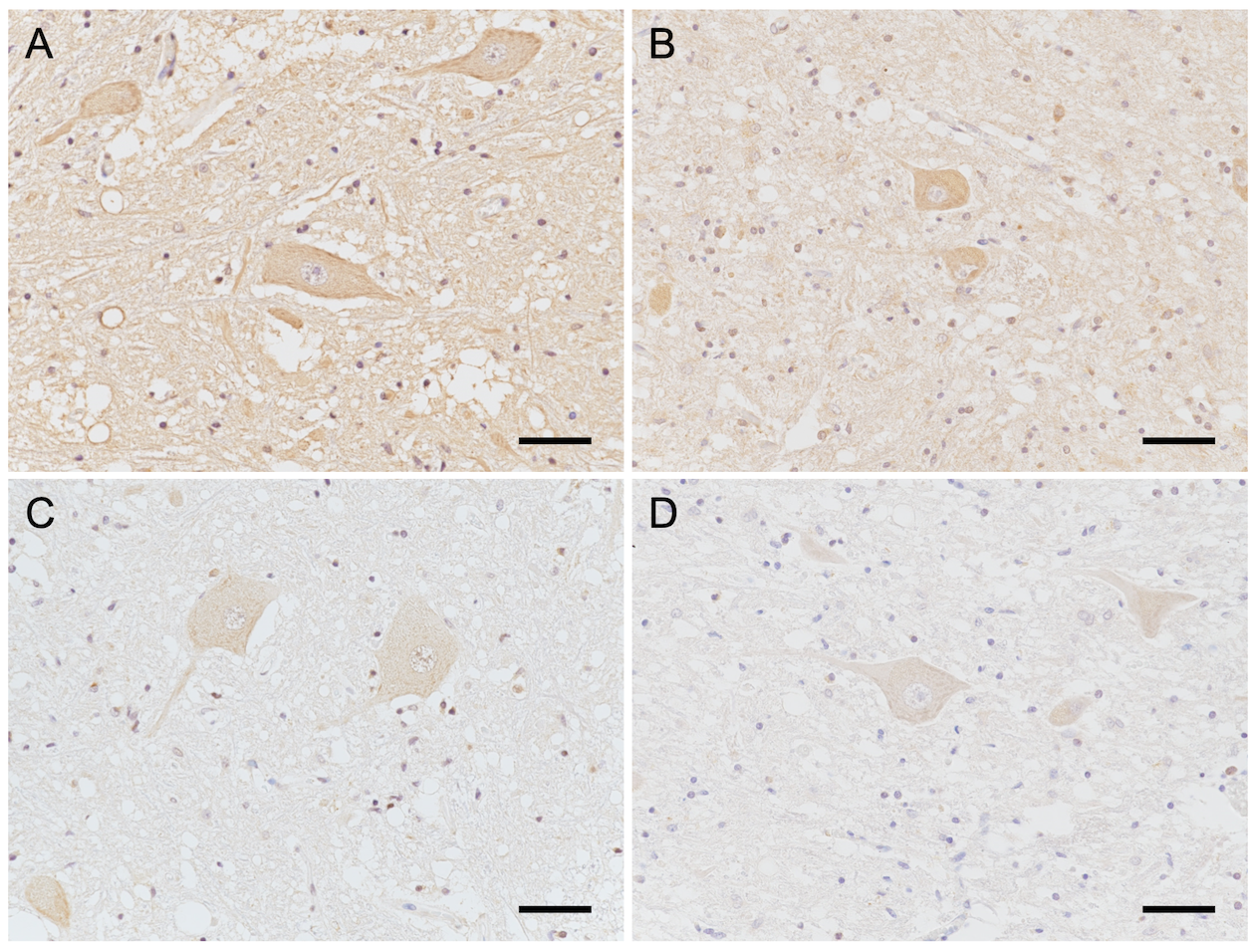
